# Biopsychosocial determinants of HPV vaccine perception in university students of both sexes in Cúcuta, Colombia, 2024: a cross-sectional study

**DOI:** 10.64898/2026.06.18.26356011

**Authors:** Andrés Arias-Sánchez, Martha Flórez, Néstor Pereira, Dixon Yirvaldo Cáceres Peñaloza, Linny Sharick Dallos Rivera, Katherine Guiselle Núñez Villamizar, Claudia Beltrán Arroyave

## Abstract

Colombia has been internationally recognised as a paradigmatic case of vaccine confidence crisis since the 2014 Carmen de Bolívar event, and national HPV vaccination coverage remains far below the World Health Organization 2030 target. Most published evidence focuses on female adolescents and on cervical cancer; the perception of the HPV vaccine in university-age populations of both sexes—and across the broader spectrum of HPV-attributable disease—remains comparatively understudied. We aimed to describe the influence of biopsychosocial determinants on HPV vaccine perception among university students of both sexes in Cúcuta, Norte de Santander, Colombia. We conducted a cross-sectional study with a mixed quantitative–qualitative approach in 2024 among four universities (Universidad de Santander, Universidad Francisco de Paula Santander, Universidad de Pamplona and Universidad Libre; combined enrolment 21,033 students). Using convenience sampling stratified by institution, 750 actively enrolled undergraduate students of both sexes (18–60 years) completed a structured online questionnaire adapted from previously validated instruments. The instrument captured sociodemographic information, HPV knowledge and HPV vaccine perception. Data were analysed using Student’s t-test, one-way analysis of variance, Tukey post-hoc tests, effect sizes and 95% confidence intervals, with a 0.05 significance threshold. Of 750 respondents, 54.2% were women, 61.3% were under 20 years of age, and 75.1% attended public universities. HPV knowledge was high in 39.2%, intermediate in 42.4% and low in 18.4%; women and students aged 26 years or older displayed higher knowledge. Although 91.2% had heard of HPV and 82.5% knew that both sexes could acquire it, recognition of clinical manifestations and complications was uneven: cervical cancer 51.7%, penile cancer 30.5%, vaginal warts 45.9% and warts in the penis, larynx, anus or rectum 34.0%. Vaccine-specific knowledge was low in 77.1%, with men disproportionately represented (85.9% versus 69.5% in women). Overall positive perception of HPV vaccination was 66.6%, slightly higher in women (68.8%) than men (63.9%), in students aged 26 years or older (70.1%) and in students from private universities (68.1% versus 65.9%). Inferential analysis identified sex (Cohen’s d = −0.357), type of university (d = 0.189) and HPV knowledge (partial eta-squared = 0.096) as the only significant determinants. Age, socioeconomic stratum, age at sexual debut and vaccine-specific knowledge did not reach meaningful significance. HPV vaccine perception was predominantly positive but conditioned by three biopsychosocial determinants, with HPV knowledge as the primary driver. The persistent gender gap reflects historical anchoring of HPV messaging in cervical disease and female-targeted campaigns. Public-health strategies should adopt comprehensive, gender-inclusive educational interventions that explicitly visibilise non-cervical HPV-related cancers and address both sexes from a common evidence base.

**Author summary:** Vaccination against the human papillomavirus prevents not only cervical cancer in women but also cancers of the mouth and throat, anus and penis, as well as anogenital warts in both sexes. In Colombia, vaccine acceptance collapsed after a 2014 episode in El Carmen de Bolívar, in which adolescent girls reported symptoms after immunization. Although the symptoms were not caused by the vaccine, public confidence has not fully recovered and most communication continues to frame the vaccine as a protection against cervical cancer in women only. We surveyed 750 university students of both sexes in Cúcuta, Colombia, to understand how they perceive the vaccine and which personal and social factors shape that perception. We found that most students view the vaccine positively, but their perception depends mainly on how much they know about the human papillomavirus itself—particularly about diseases beyond cervical cancer. Men knew less than women and were less aware of the diseases that affect men. Type of university also played a small role. Our results suggest that improving HPV education for both sexes, and explicitly explaining that the vaccine prevents disease in men too, would help Colombia restore confidence and reach the World Health Organization 2030 vaccination targets.

## Introduction

Human papillomavirus (HPV) is among the most prevalent sexually transmitted infections worldwide, with an estimated global prevalence of approximately 11.7%; most sexually active individuals acquire the infection at some point in life [1,2]. More than 200 HPV serotypes have been described, of which around 40 affect mucosal surfaces and are classified into high- and low-risk types. High-risk genotypes, particularly HPV-16 and HPV-18, are causally associated with cervical, anal, penile and oropharyngeal cancers, making HPV a public-health problem that affects both sexes [3].

Prophylactic HPV vaccination is one of the most effective preventive tools available against this group of cancers. Colombia introduced the quadrivalent vaccine in the Expanded Programme on Immunization in 2012, achieving first-dose coverage above 95% in the initial cohorts. In 2014, however, an event in El Carmen de Bolívar—interpreted by the National Institute of Health and the Pan American Health Organization as mass psychogenic illness without organic substrate—precipitated an abrupt collapse of public confidence, with documented reductions of approximately 14% in first dose and 5% in completion [4,5]. The COVID-19 pandemic subsequently caused additional disruption, with declines between 16.1% and 39.4% in first dose and between 3.4% and 11.8% in second dose during 2021 [6].

Vaccine perception is shaped by social, cultural and personal factors that vary by context, time, place and vaccine type [7,8]. Biopsychosocial determinants—age, previous experiences, side effects of other vaccines, knowledge about the virus and its relationship with cancer, religious beliefs and trusted communication channels—have been recurrently identified as drivers of HPV vaccine acceptance and attitudes [9–11]. University students, at a transitional life stage and as future health promoters in their families and communities, are a particularly relevant population in which to study perception. Their attitudes can be positive, anchored in recognition of the vaccine’s safety and efficacy, or negative, marked by fear of adverse events, misinformation and social taboos.

Inequitable perception translates into uneven coverage, more HPV infections and a higher incidence of cervical and other HPV-related cancers. In Colombia, 24,689 prevalent cervical cancer cases were registered in 2020, rising to 30,997 in 2022—a 17% increase [12,13]. Importantly, public-health communication has historically framed HPV vaccination almost exclusively around cervical cancer, contributing to a gendered perception of the vaccine and obscuring the substantial burden of HPV-attributable disease in men (oropharyngeal, anal and penile cancers, as well as anogenital warts).

Despite the relevance of this issue, the perception of the HPV vaccine in Colombian university populations of both sexes has been comparatively understudied, and the specific role of biopsychosocial determinants in this group remains incompletely characterised. We therefore conducted this study to describe the influence of biopsychosocial factors on HPV vaccine perception in a large sample of university students of both sexes in Cúcuta, Norte de Santander, during 2024, with the explicit aim of informing the design of comprehensive, gender-inclusive educational and communication strategies.

## Materials and methods

### Study design and setting

Cross-sectional study with a mixed quantitative–qualitative approach, conducted during 2024 in the metropolitan area of Cúcuta, Norte de Santander, Colombia. Four universities were included: Universidad de Santander (UDES), Universidad Francisco de Paula Santander (UFPS), Universidad de Pamplona and Universidad Libre, with a combined enrolment of 21,033 students. Reporting follows the Strengthening the Reporting of Observational Studies in Epidemiology (STROBE) statement for cross-sectional studies; the completed checklist is provided as Supporting Information (S1 Checklist).

### Participants and sampling

Eligible participants were undergraduate students of both sexes, all aged 18 to 60 years, none minors for legally adults in Colombia and actively enrolled in the academic period of 2024. Recruitment used a convenience sampling strategy stratified by institution, distributing the sample as follows: 124 students from UDES, 263 from Universidad de Pamplona, 251 from UFPS and 112 from Universidad Libre, for a total of 750 valid responses. This sample size yields a maximum margin of error of approximately ±3.6% for population proportions at 95% confidence in the combined target population, allowing meaningful subgroup analyses by sex, age and type of institution.

### Data collection instrument

A structured questionnaire was designed by adapting and triangulating items from previously published instruments: the Puerto Rico Institute of Statistics HPV Survey [14], Lema-Vera and colleagues [15], Frietze and colleagues [16], and the DANE Survey of Culture and Politics [17]. The instrument comprised three sections: (i) sociodemographic and socioeconomic information; (ii) HPV knowledge, including modes of transmission, prevention and association with HPV-attributable conditions across both sexes; and (iii) perception of the HPV vaccine, including dimensions of trust, perceived benefit, perceived quality, perceived inconveniences and willingness to recommend. The instrument is provided as Supporting Information (S1 Questionnaire).

The instrument was pilot-tested on a small group of students (n = 20) to assess face validity, clarity and time to completion; minor wording adjustments were made before deployment. The final version was distributed virtually via Google Forms, ensuring anonymity, voluntary participation and informed consent before data entry [18].

### Variables and operational definitions

HPV knowledge was scored on a 10-point composite scale based on items covering transmission, prevention and clinical manifestations; categories were defined a priori as low (≤5), intermediate (6–7) and high (≥8). Vaccine knowledge was scored similarly on items addressing schedule, target population and effectiveness. Perception was operationalised as a multi-item Likert scale and grouped into fully positive perception, mostly positive perception, neutral, mostly negative and fully negative. Biopsychosocial covariates included sex, age, socioeconomic stratum (Colombian SISBEN classification), age at sexual debut, type of university (public versus private), HPV knowledge and vaccine knowledge.

### Statistical analysis

Data were managed in Microsoft Excel and analysed in IBM SPSS Statistics. Descriptive analysis used central-tendency measures (mean, median, mode) and frequency distributions. Inferential analysis used Student’s t-test for two-group comparisons (sex, type of university) and one-way analysis of variance (ANOVA) for three-group comparisons (age groups, knowledge categories, socioeconomic strata). Levene’s test was used to assess homogeneity of variances. Effect size was reported as Cohen’s d for t-tests and partial eta-squared for ANOVA, with 95% confidence intervals. Statistical power was estimated for the main contrasts. Post-hoc comparisons used Tukey’s honest significant difference test. The significance threshold was set at 0.05.

### Ethics statement

The study was conducted in accordance with the Declaration of Helsinki and Colombian Resolution 8430 of 1993 of the Ministry of Health, which classifies survey-based studies of this nature as minimal-risk research. Ethical approval was obtained from the Research Ethics Committee of Hospital Universitario Erasmo Meoz (approval code 2024-136-015072-2). All participants provided informed consent electronically before responding; participation was anonymous and voluntary, with no personally identifiable information collected. De-identified data are stored in a password-protected repository accessible only to the research team.

## Results

### Sociodemographic profile

Of 750 valid responses, 54.2% (n = 407) were women; 61.3% (n = 419) were under 20 years of age and 41.5% (n = 284) were between 21 and 25 years; 75.1% (n = 514) attended public universities.

### HPV knowledge

Overall HPV knowledge was high in 39.2% (n = 294), intermediate in 42.4% (n = 318) and low in 18.4% (n = 138). Women showed higher mean scores than men (7.13 versus 6.24), and the proportion of high knowledge was greater in students aged 26 years or older (59.6%). Detailed distributions by sex, age and type of university are shown in Table 1.

**Table 1.** Distribution of HPV knowledge among university students of Cúcuta, 2024 (n = 750).

| Group/subgroup | Mean score | Low (n) | Low % | Intermediate (n) | Intermediate % | High (n) | High % |
| --- | --- | --- | --- | --- | --- | --- | --- |
| **General** | 6.728 | 138 | 18.4 | 318 | 42.4 | 294 | 39.2 |
| **Women** | 7.13 | 50 | 12.3 | 179 | 44.0 | 178 | 43.7 |
| **Men** | 6.24 | 88 | 25.7 | 139 | 40.6 | 115 | 33.6 |
| **Age <20** | 6.276 | 79 | 18.9 | 178 | 42.5 | 162 | 38.7 |
| **Age 21–25** | 6.721 | 52 | 18.3 | 128 | 45.1 | 104 | 36.6 |
| **Age >26** | 7.659 | 71 | 14.9 | 2 | 25.5 | 28 | 59.6 |
| **Private university** | 6.813 | 43 | 18.2 | 92 | 39.0 | 101 | 42.8 |
| **Public university** | 6.688 | 95 | 18.5 | 226 | 44.0 | 193 | 37.5 |

Awareness of basic HPV facts was generally high: 91.2% (n = 684) had heard of HPV, 87.6% (n = 657) identified it as a sexually transmitted infection, 82.5% (n = 619) recognised that both men and women can acquire it, and 69.7% (n = 522) correctly identified routes of transmission. Among preventive methods, 78.3% recognised condom use as a primary measure, 76.1% (n = 571) recognised vaccination and only 24.8% (n = 186) identified abstinence. Recognition of HPV-attributable conditions was uneven: cervical cancer 51.7% (n = 388), penile cancer 30.5% (n = 229), vaginal warts 45.9%, and warts in the penis, larynx, anus or rectum 34.0%. Notably, only 38.9% of those identifying cervical cancer as a complication were men, and only 26% of those identifying warts in the penis, oropharynx, anus or rectum were men, reflecting a clear gendered asymmetry in recognition of HPV-related disease.

### HPV vaccine knowledge

Overall vaccine-specific knowledge was low: 77.1% (n = 578) of respondents had low knowledge, 19.3% (n = 145) intermediate and only 3.6% (n = 27) high. Men were disproportionately represented in the low-knowledge category (85.9%; n = 294 of male respondents) compared with women (69.5%; n = 283). The same pattern was observed across items: women and students under 20 years had the highest knowledge levels.

### Overall perception of HPV vaccination

Sixty-six point six percent of students held a positive perception of HPV vaccination. By sex, women displayed slightly higher proportions of positive perception (68.8%) than men (63.9%). By age, students aged 26 years or older reached the highest positive perception (70.1%), followed by 21–25 years (67.3%) and under 20 years (65.5%). By type of university, private-institution students reached 68.1% positive perception versus 65.9% in public institutions.

### Biopsychosocial determinants of perception

Inferential analysis identified sex, type of university and HPV knowledge as the only statistically significant determinants of perception. Effects on perception by age, socioeconomic stratum, age at sexual debut and vaccine-specific knowledge did not reach meaningful significance with relevant effect sizes (Table 2).

**Table 2.**
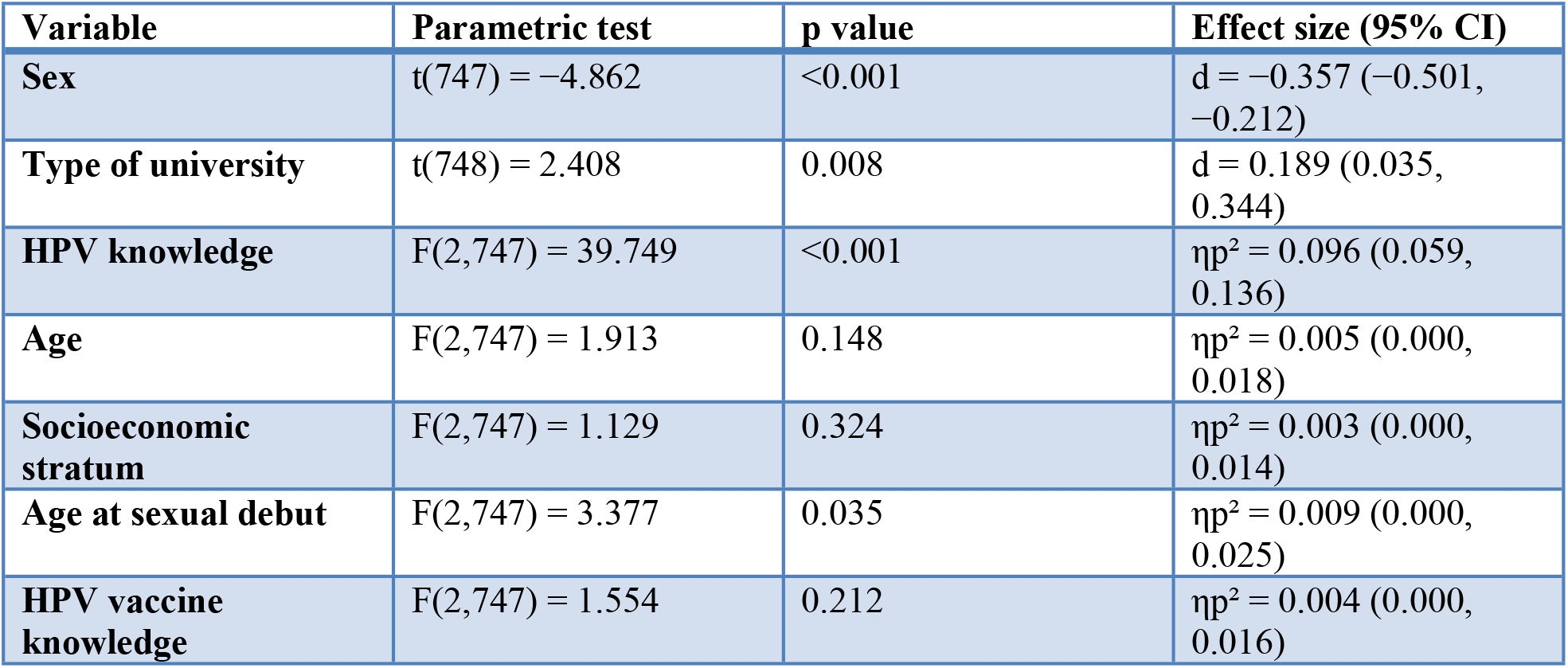
Inferential analysis of biopsychosocial determinants of HPV vaccine perception (n = 750).

## Discussion

Among 750 university students of both sexes in Cúcuta in 2024, HPV vaccine perception was predominantly positive (66.6%) and was significantly shaped by three biopsychosocial determinants: HPV knowledge, sex and type of university. Knowledge about HPV—not knowledge about the vaccine— emerged as the largest single contributor (partial eta-squared = 0.096; approximately 10% of explained variance), with sex contributing a small-to-moderate effect (Cohen’s d = −0.357) and type of university a small effect (d = 0.189). The determinants form a hierarchy: type of university appears to operate indirectly, through differences in HPV knowledge, rather than as an independent driver.

Our finding that 91.2% of students had heard of HPV is consistent with previously reported figures of around 80% in young adults of African American and African immigrant origin [9]. However, awareness does not translate automatically into understanding: 82.5% of our respondents knew that both sexes can be infected, but only half could link HPV to specific conditions such as cervical cancer, penile cancer or anogenital warts. This dissociation between general awareness and specific disease recognition has been described internationally and is a recurrent target of HPV educational interventions [8,11,16].

The gendered asymmetry of disease recognition in our sample is particularly informative. Men were under-represented among those who identified cervical cancer (38.9% of recognisers) and even more so among those who identified warts of the penis, oropharynx, anus and rectum (26%). This pattern is consistent with previous findings that the public perceives the HPV vaccine as more related to women than to men, based on the belief that aggressive cancers occur exclusively in women and that men only transmit the disease [24]. Our results confirm this gendered cognitive structure in a Colombian university population and reinforce the need for a comprehensive, gender-inclusive narrative.

Positive aspects of perception—trust in the vaccine (39.7%), low perceived inconveniences (57.87%) and willingness to recommend (92.53%)—are consistent with previous evidence that perceived benefits explain a substantial increase in intention to vaccinate children once knowledge is controlled for. Negative perception drivers—fear of adverse events, fear of secondary disease, suspicion of scientific experimentation and risk-benefit uncertainty—are likewise consistent with the determinants described by the Pan American Health Organization during the COVID-19 vaccination campaign [22] and by previous parental perception studies in the Andean region [23].

Our findings carry two practical implications. First, since HPV knowledge is the dominant determinant of perception, educational interventions are likely to be more effective than purely promotional ones. Specifically, interventions should foreground the multi-site, multi-sex profile of HPV-attributable disease—oropharyngeal, anal, penile, vulvar, vaginal and cervical cancers, alongside anogenital warts—rather than relying on the cervical cancer narrative that has historically dominated Colombian campaigns. Second, the persistent gender gap suggests that male-specific communication is urgently needed; the recent expansion of the Colombian HPV vaccine schedule to boys (2023) and adolescents up to 17 years (2024) [20] provides an opportunity, but its success depends on a parallel cultural shift in messaging.

### Strengths and limitations

Strengths include a relatively large sample (n = 750) drawn from four distinct universities, the simultaneous inclusion of men and women, the use of validated source instruments, and inferential analyses with explicit effect sizes and confidence intervals. The study also reports against the STROBE checklist for cross-sectional studies.

Limitations should be acknowledged. First, convenience sampling restricts external validity, although stratification by institution mitigated selection imbalances; results may not generalise beyond Cúcuta or to non-university youth. Second, self-reported online responses are subject to social-desirability bias and self-selection. Third, the cross-sectional design precludes causal inference. Fourth, the relatively low effect sizes for sex and type of university imply that, although statistically significant, their individual practical contributions are modest compared with knowledge. Fifth, the instrument captured general perception dimensions; future studies should incorporate validated multi-dimensional scales. Sixth, recognition of clinical manifestations was assessed at the level of awareness rather than depth of understanding; qualitative work should complement these quantitative findings.

## Conclusions

Among university students of both sexes in Cúcuta in 2024, HPV vaccine perception was predominantly positive but conditioned by HPV knowledge (primary determinant), sex and type of university (secondary determinants operating largely through knowledge). The persistent gender gap reflects the historical anchoring of HPV messaging in cervical cancer and the female-targeted design of early campaigns. Public-health policy should adopt a comprehensive, gender-inclusive communication framework that explicitly visibilises non-cervical HPV-related conditions and addresses both sexes from a common evidence base. Investments in deep, knowledge-focused educational interventions are more likely to be effective than promotional campaigns alone.

## Data Availability

Yes, data can be available if needed

## Acknowledgments

The authors thank the participating students of Universidad de Santander, Universidad Francisco de Paula Santander, Universidad de Pamplona and Universidad Libre, as well as the academic staff who facilitated questionnaire distribution. We also thank the Research Ethics Committee of the Hospital Universitario Erasmo Meoz for the timely review and approval of the study protocol.

## Supporting information

S1 Checklist. STROBE checklist for cross-sectional studies.

S1 Questionnaire. Structured questionnaire used in the study (Spanish original and English translation).

S1 Dataset. De-identified individual-level dataset (available from the corresponding author upon reasonable request, subject to ethical and institutional approval).

## Notes

### Competing Interest Statement

The authors have declared no competing interest.

### Funding Statement

No financial funding was required

### Author Declarations

Ethical approval was obtained from the Research Ethics Committee of Hospital Universitario Erasmo Meoz (approval code 2024-136-015072-2).

## References

1. Munive JE, Gamboa PS, González MA, González JS. Rapid review: HPV vaccination in children and men. Rev Salud Bosque. 2024;14(1):1–13.

2. World Health Organization. Human papillomavirus and cancer. Geneva: WHO; 2022. Available from: https://www.who.int/news-room/fact-sheets/detail/human-papilloma-virus-and-cancer

3. Toro-Montoya AI, Tapia-Vela LJ. Human papillomavirus (HPV) and cancer. Medicina & Laboratorio. 2022.

4. Palencia-Sánchez F, Echeverry-Coral SJ. Social aspects affecting acceptance of HPV vaccination in Colombia: a systematic review. Rev Colomb Obstet Ginecol. 2020;71(2):178–194. doi:10.18597/rcog.3448

5. Simas C, Muñoz N, Arregoces L, Larson HJ. HPV vaccine confidence and cases of mass psychogenic illness following immunization in Carmen de Bolívar, Colombia. Hum Vaccin Immunother. 2019;15(1):163–166. doi:10.1080/21645515.2018.1511667

6. El Espectador. Who will save HPV vaccination in Colombia? [Online]. 15 December 2021.

7. Mayoral LE, Hapon MB, Troncoso M, Miras DN, Romero GS. Vaccines and immunity: a didactic study based on secondary-school students’ perceptions in Mendoza, Argentina. Eur Sci J. 2020;16(18):24.

8. Larson HJ, Jarrett C, Eckersberger E, Smith D, Paterson P. Understanding vaccine hesitancy around vaccines and vaccination from a global perspective: A systematic review of published literature, 2007–2012. Vaccine. 2014;32(19):2150–2159.

9. Adegboyega A, Obielodan O, Wiggins AT, Dignan M, Williams LB. Beliefs and knowledge related to HPV vaccine among African Americans and African immigrants young adults. Cancer Causes Control. 2023;34(5):479–489.

10. Shen F, Du Y, Cao K, Chen C, Yang M, Yan R, et al. Acceptance of the human papillomavirus vaccine among general men and men with a same-sex orientation and its influencing factors: A systematic review and meta-analysis. Vaccines (Basel). 2023;12(1):16.

11. Frietze G, Padilla M, Cordero J, Gosselink K, Moya E. Human Papillomavirus Vaccine Acceptance (HPV-VA) and Vaccine Uptake (HPV-VU): assessing the impact of theory, culture, and trusted sources of information in a Hispanic community. BMC Public Health. 2023;23(1).

12. UNICEF. The COVID-19 pandemic has caused the largest backslide in vaccination in 30 years. New York: UNICEF; 2022.

13. Cuenta de Alto Costo. World Cervical Cancer Day 2022. Bogotá: CAC; 2022.

14. Puerto Rico Institute of Statistics. Survey on Human Papillomavirus (HPV) in Adults. San Juan: PRIS; 2013.

15. Lema-Vera LA, Mesa-Cano IC, Ramírez-Coronel AA, Jaya-Vásquez LC. Knowledge about HPV in upper-basic and high-school students. 2021.

16. Frietze G, Padilla M, Cordero J, Gosselink K, Moya E. HPV-VA and HPV-VU in a Hispanic community. BMC Public Health. 2023;23(1).

17. Departamento Administrativo Nacional de Estadística (DANE). Survey of Political Culture – ECP. Bogotá: DANE; 2023.

18. Bustamante-Ramos GM, Martínez-Sánchez A, Tenahua-Quitl I, Jiménez C, López-Mendoza Y. Knowledge and prevention practices about HPV in university students of the Sierra Sur, Oaxaca. An Fac Med. 2015;76(4):369– 376.

19. Instituto Departamental de Salud. Bulletin on cervical cancer P2 2023. Norte de Santander: IDS; 2023.

20. Ministerio de Salud y Protección Social (Colombia). National vaccination campaign 2025. Bogotá: MinSalud; 2025.

21. Ossa CA, Torres D, Rojas P, Cuello M, Recabal P, Quezada N, et al. Characterisation of breast cancer patients under 40 years of age in a university hospital. Rev Chil Obstet Ginecol. 2014;79(6):373–379.

22. Pan American Health Organization. Misinformation fuels doubts about COVID-19 vaccines, says PAHO director. Washington: PAHO; 2021.

23. Cano Ugarte B, Hidalgo Zarate CJ. Perceptions and knowledge of HPV and its vaccination among parents in the municipality of Cliza. Rev Cient Enferm UNITEPC. 2023;4(1):14–20.

24. Schwartz BI, Maccani M, Bansal S, Gannon M. Parental perceptions of the HPV vaccine for prevention of anogenital and oropharyngeal cancers. Vaccine X. 2023;14:100298.

